# Fetomaternal outcomes among cesarean section parturients administered with norepinephrine vs. phenylephrine for post-spinal anesthesia hypotension: A systematic review and meta-analysis

**DOI:** 10.1101/2025.03.04.25323394

**Authors:** Alyssa Camba, Rishi Denice S. Ganadin, Amador F. Simando, Paul K. Yu, Jowi Tsidkenu P. Cruz

## Abstract

Post-spinal anesthesia hypotension during cesarean delivery poses significant risks to maternal cardiac output and fetal oxygenation. Although phenylephrine (PE) is the standard vasopressor, its use is linked to an increased incidence of maternal bradycardia. In contrast, norepinephrine (NE) offers a more favorable hemodynamic profile and is emerging as a promising alternative. This systematic review and meta-analysis of 18 randomized controlled trials compared NE and PE for the prevention and treatment of post-spinal hypotension in cesarean section parturients. Our findings demonstrate that while NE and PE are equally effective in managing hypotension, NE significantly reduces the incidence of maternal bradycardia (OR = 0.49 [CI: 0.38 to 0.62]) and shows a trend toward fewer adverse maternal events, such as dizziness and reactive hypertension. Additionally, neonatal outcomes indicated a lower birth weight with NE—although still within the normal range—along with tendencies toward lower umbilical arterial lactate levels and improved umbilical vein blood gas pH. These results support NE as a viable alternative to PE, particularly for lowering the incidence of maternal bradycardia, and provide crucial evidence for updating clinical practice guidelines to enhance maternal and neonatal care during cesarean deliveries.

**Systematic review registration ID: CRD42024593459:** https://www.crd.york.ac.uk/PROSPERO/view/CRD42024593459

## Introduction

The World Health Organization (WHO, 2021) reported that 95% of all maternal deaths occur in low- and lower-middle-income countries, with nearly 800 women dying daily from preventable causes related to pregnancy and childbirth. One such preventable cause is hypotension, defined as abnormally low blood pressure, typically a systolic blood pressure of less than 90 mmHg. In cesarean section (CS) parturients, post-spinal anesthesia hypotension occurs primarily due to a decrease in total peripheral resistance (TPR) caused by sympathetic blockade, leading to vasodilation. This reduction in TPR results in decreased venous return which in turn lowers stroke volume and cardiac output, ultimately contributing to the drop in blood pressure (Sharma et al., 2021). Recent studies estimate the incidence of post-spinal anesthesia hypotension in CS deliveries to range from 1.7% to 71% (Karnina et al., 2022; Munyanziza, 2022; Shitemaw et al., 2020; Butwick et al., 2015), while Herbosa et al. (2022) report rates as high as 80% to 90% in Southeast Asia. Despite this variation, it is evident that hypotension following anesthesia administration is a significant concern during CS deliveries due to its potential to reduce maternal cardiac output and compromise fetal oxygenation (Šklebar et al., 2019).

Phenylephrine (PE) is favored in the consensus statement as the primary vasopressor for managing hypotension after the administration of spinal anesthesia for cesarean deliveries in parturients with normal heart rates (Herbosa et al., 2022). Concurrently, norepinephrine (NE) is considered as an alternative option for addressing hypotension and bradycardia based on Randomized Controlled Trials (RCTs) across existing studies that have proven its efficacy albeit the need to expand its credibility is critical–as its use in post-spinal hypotension is relatively new, and literature on the topic remains scarce (Herbosa et al., 2022). While previous meta-analyses have compared the use of PE and NE (Kumari et al., 2022; Liu et al., 2022), uncertainties persist regarding their comparative effects on maternal hemodynamic stability and neonatal outcomes, including maternal blood pressure, heart rate, and the potential for bradycardia during cesarean section (Stewart et al., 2010).

PE-induced hemodynamic changes, such as reflex bradycardia, can reduce maternal cardiac output and pose risks in cases of fetal acidosis, particularly during emergency cesarean sections (Kalra and Hayaran, 2011; Stewart et al., 2010). While a decrease in heart rate may not adversely affect the fetus in elective surgeries, the same effect can exacerbate fetal compromise in urgent scenarios (Kalmar et al., 2018). Maternal bradycardia resulting from PE use can influence neonatal outcomes, although its overall impact on the mother and the neonate varies. For instance, a meta-analysis comparing norepinephrine and phenylephrine found that while norepinephrine was associated with a lower incidence of maternal bradycardia, there were no significant differences in neonatal APGAR scores or umbilical vein blood gas values between the two vasopressors (Xu et al., 2019). Studies indicate that PE, compared to other vasopressors such as ephedrine, is associated with lower incidences of fetal acidosis (Cooper et al., 2002) but an increased incidence of maternal bradycardia (Xu et al., 2018). While direct evidence linking maternal bradycardia to reduced placental perfusion is limited, related studies suggest that maternal cardiovascular changes can impact placental blood flow and fetal well-being, highlighting the need to balance maternal blood pressure stabilization with potential fetal risks if placental perfusion is compromised (Clark et al., 2023). Furthermore, other neonatal outcomes, including Apgar scores, the need for NICU admission, and umbilical cord blood pH levels, are critical for assessing the overall impact of vasopressors on the fetus.

The varying effects of vasopressors on both maternal and neonatal health underscore the need for comprehensive research to determine the most effective drug for preventing and managing post-spinal anesthesia hypotension. To respond to this research gap, our study aims to systematically evaluate maternal hemodynamic abnormalities and neonatal outcomes as a result of norepinephrine versus phenylephrine use in cesarean section parturients undergoing spinal anesthesia. By synthesizing evidence from randomized controlled trials, we seek to identify the optimal vasopressor for preventing or treating post-spinal hypotension, potentially guiding future updates in clinical protocols and ultimately enhancing maternal and neonatal care.

## Methods

### Literature Search

Prior to conducting the study, it was granted an “Exempt from Review” status from the Research Ethics Review Committee of De La Salle University. This systematic review and meta-analysis adhered to the Preferred Reporting Items for Systematic Reviews and Meta-Analyses (PRISMA) guidelines and the Cochrane Handbook for Systematic Reviews of Interventions (Higgins et al., 2024; Moher et al., 2009). The entire workflow is depicted in the PRISMA chart shown in Figure 1. In line with PRISMA guidelines, the study began with a clearly defined research question and protocol development, which was registered on PROSPERO under registration number <u>CRD42024593459</u>, to ensure transparency and accountability. We applied predefined eligibility criteria to select relevant studies, including only randomized controlled trials that compared norepinephrine and phenylephrine in cesarean section deliveries for the prevention or treatment of post-spinal hypotension. A systematic search was performed in PubMed and Scopus using the search strategy: (“cesarean section” OR “caesarean section” OR ”CS” OR “C section”) AND (“spinal anesthesia“ OR “spinal anaesthesia” OR “spinal block”) AND (“norepinephrine” OR “noradrenaline” OR “NE” OR “noradrenalin”) AND (“phenylephrine” OR “PE”). Studies were then screened based on the title, abstract, and full text, with data independently extracted by two reviewers.

**Fig. 1.**
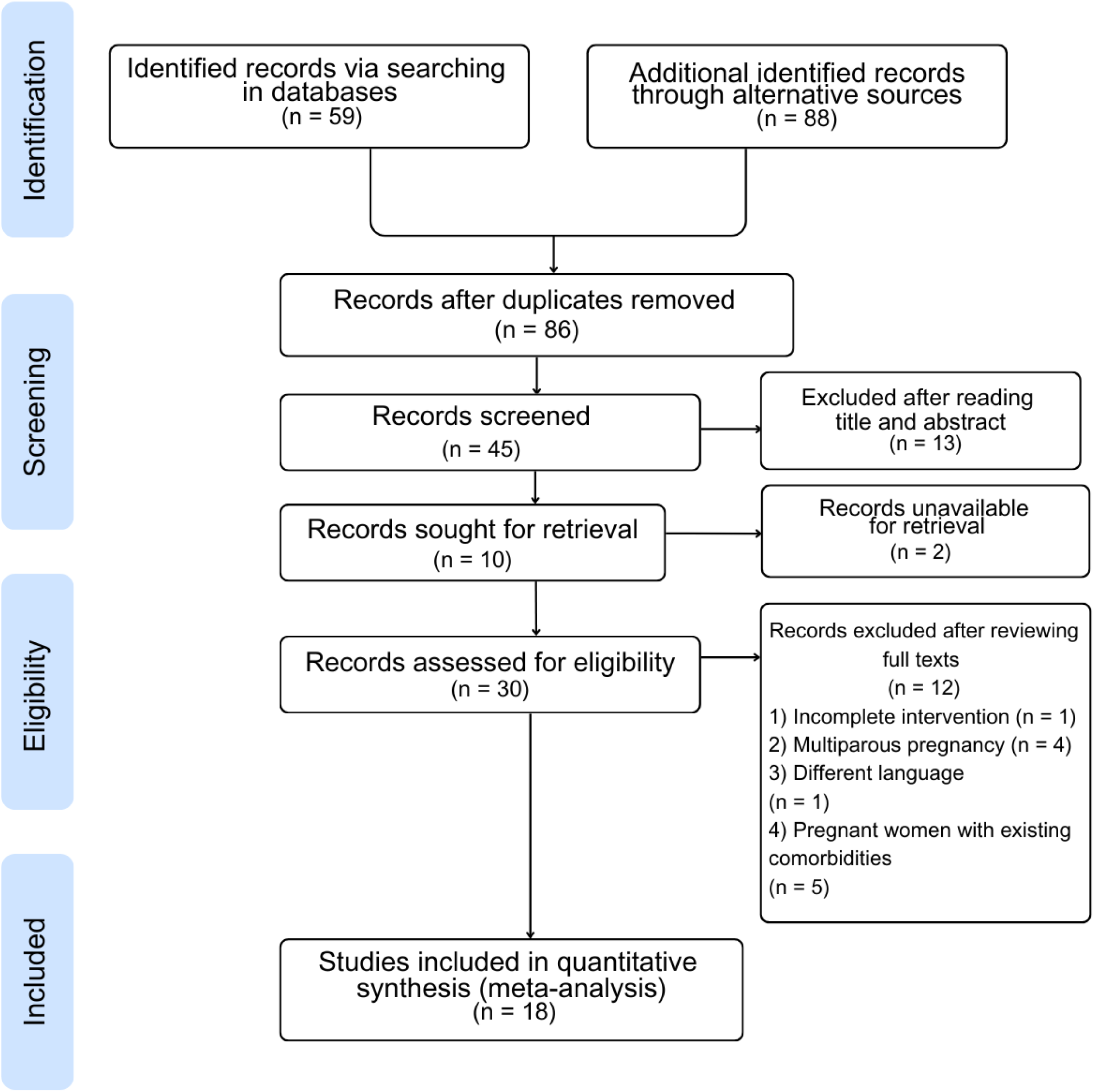
Preferred Reporting Items for Systematic Reviews and Meta-Analyses (PRISMA) chart. This chart illustrates the systematic screening and selection process, starting with 147 identified records, leading to 18 studies included in the meta-analysis after duplicates, ineligible records, and unavailable data were excluded.

### Study Selection

A total of 18 studies were included in the analysis. Eligible studies included in the current study are based on the following inclusion criteria: (a) randomized controlled trials on Phenylephrine and Norepinephrine incidence in post-spinal anesthesia hypotension, (b) study population included women that underwent cesarean section delivery, (c) the study has both norepinephrine and phenylephrine as their study of interest, (d) parameters that included maternal OR neonatal outcomes, as well as (e) identifying whether administration of vasopressors will be by infusion or bolus. The exclusion criteria on the other hand included (a) women that underwent cesarean section delivery but had multiparous, (b) the randomized controlled trial study is in a different language, and (c) pregnant women with existing comorbidities or are immunocompromised.

### Data Extraction

Two reviewers independently assessed the titles and abstracts of the studies. Full-text articles were obtained for all eligible studies and evaluated independently by three reviewers. Data irrelevant to the study were excluded, with the reasons for exclusion documented to align with the established exclusion criteria. The following data were extracted from each included study: population, intervention, comparison or control of vasopressors (phenylephrine and norepinephrine), outcomes, and study methodology (see Table 1 for details of the included studies).

**Table 1.**
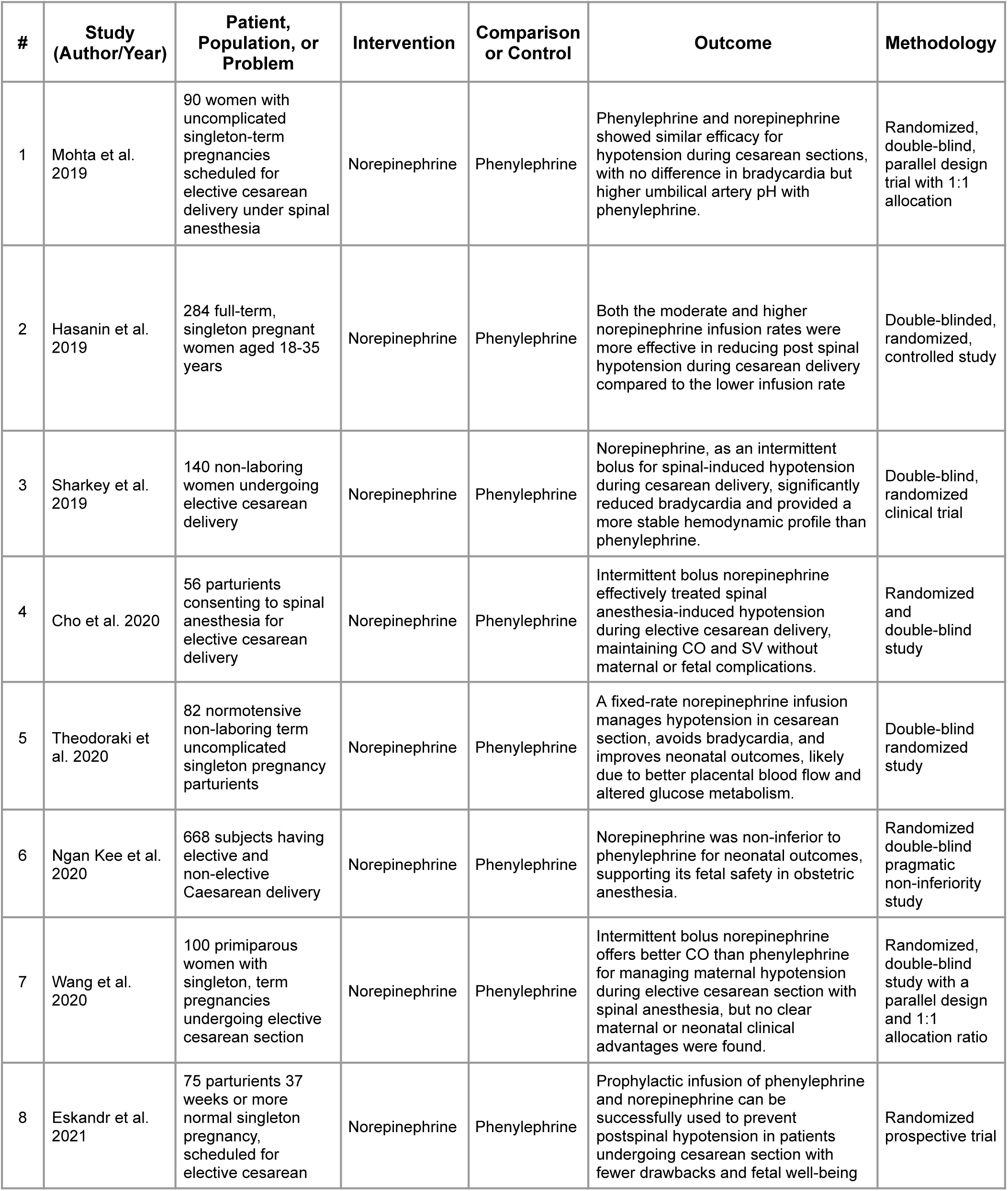
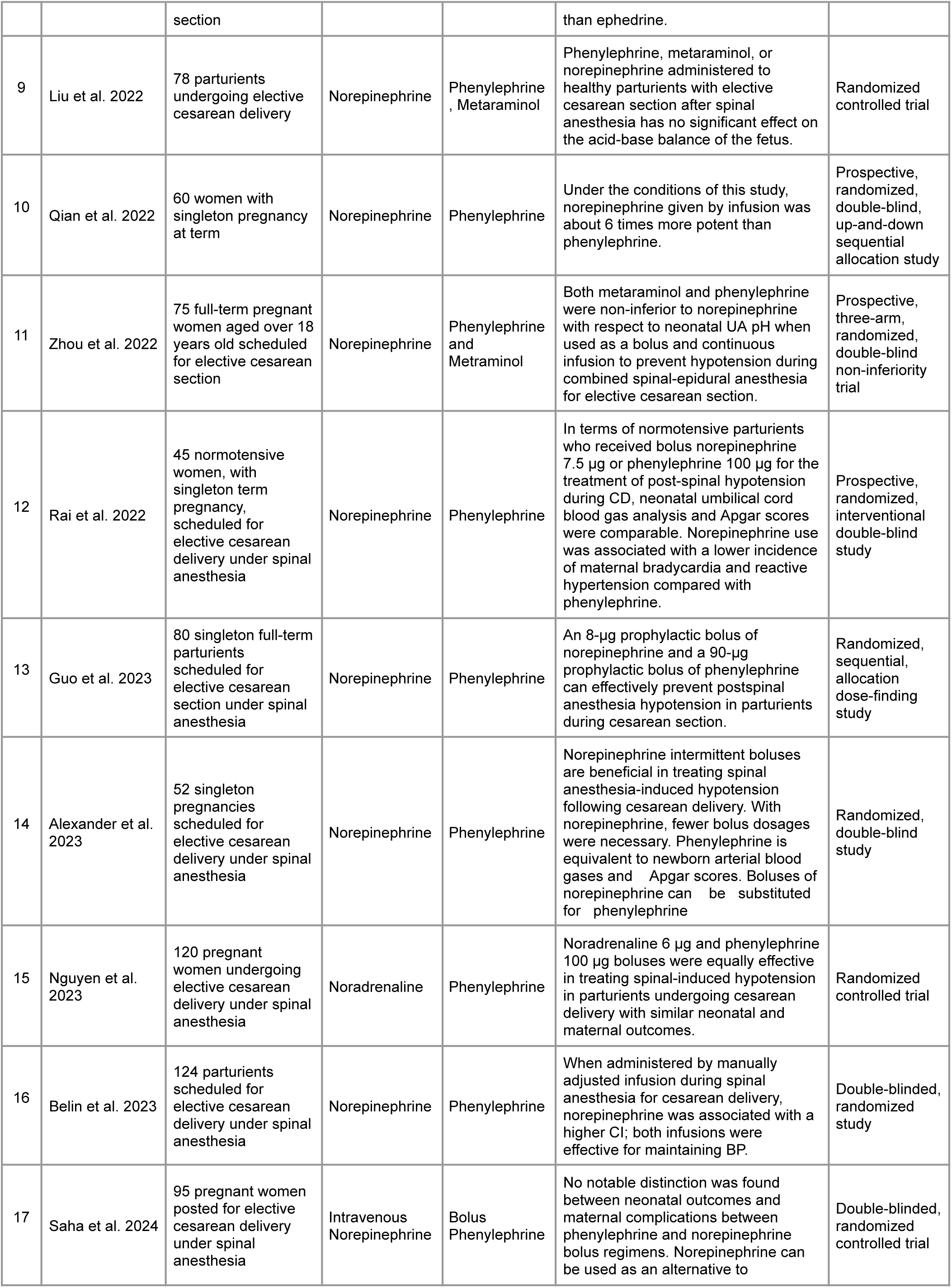
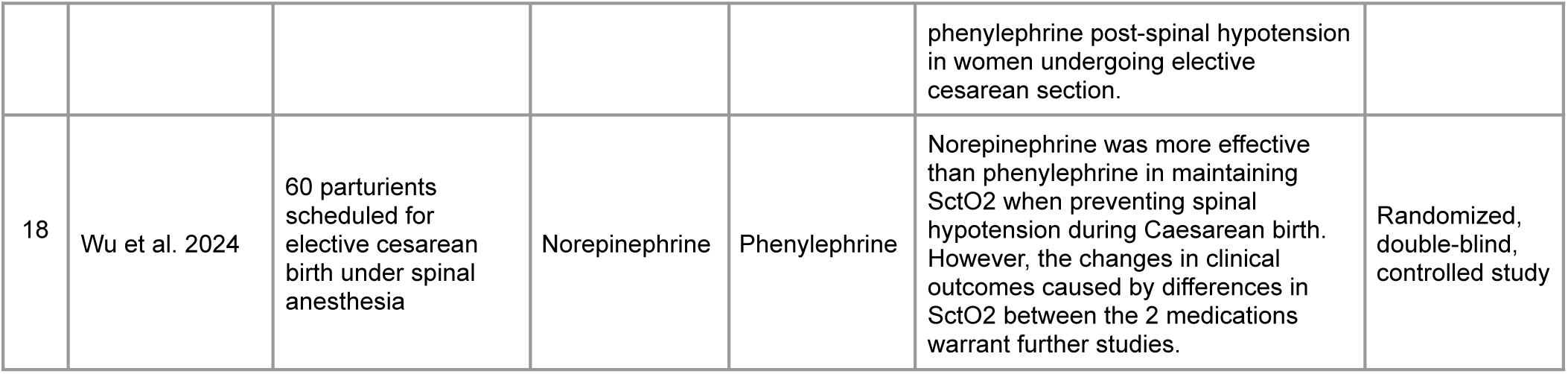
Population, intervention, comparison or control of vasopressors (phenylephrine and norepinephrine), outcomes, and study methodology (PICOM) table for the nineteen included randomized controlled trials.

### Assessment of Study Quality

The quality of the included studies was assessed using the Cochrane Risk of Bias version 2.0 (RoB2), a tool from the Cochrane Collaboration designed for evaluating the risk of bias in randomized controlled trials. The RoB2 assessment tool was applied to assess potential bias across six domains: (1) random sequence generation, (2) allocation concealment, (3) blinding of participants and personnel, (4) blinding of outcome assessment, (5) incomplete outcome data, and (6) selective reporting. (Higgins et al., 2024) The ROB (Risk of Bias) table for the nineteen included studies is shown in Figure 2.

**Fig. 2.**
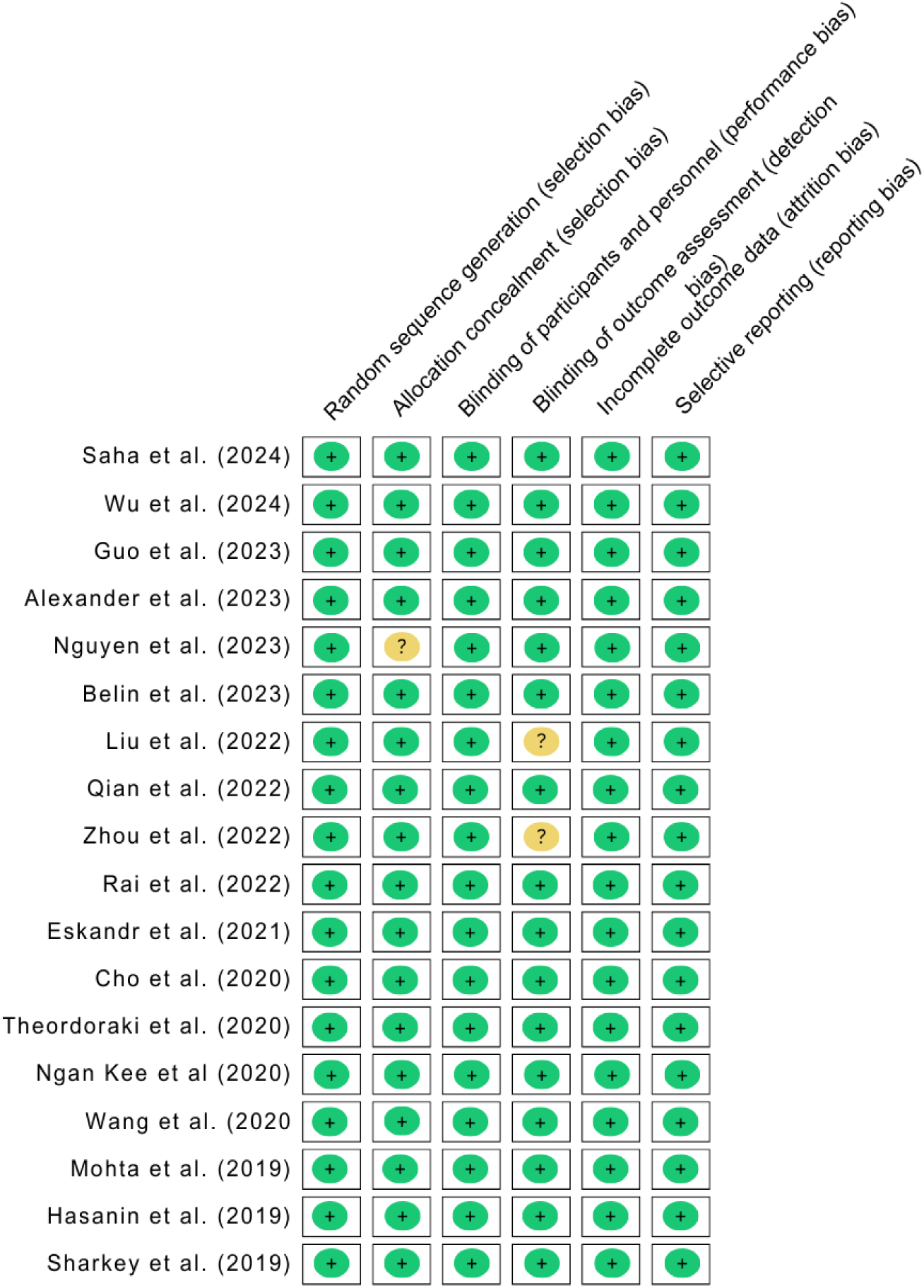
Risk of Bias (RoB) table for the nineteen included studies. Most studies are rated as having a low risk of bias (green), while three studies show a moderate risk (yellow). The Cochrane Collaboration RoB2 tool was applied to assess potential bias across six domains: (1) random sequence generation, (2) allocation concealment, (3) blinding of participants and personnel, (4) blinding of outcome assessment, (5) incomplete outcome data, and (6) selective reporting.

### Statistical Analysis

We employed forest plots generated using Review Manager (RevMan) Version 5.4 to assess the strength of associations between the administration of phenylephrine and norepinephrine and hemodynamic abnormalities and neonatal outcomes in cesarean section parturients, specifically for the prevention OR treatment of post-spinal anesthesia hypotension. The strength of these associations was expressed using odds ratios (OR) and mean differences (MD) with a 95% confidence interval.

To address potential heterogeneity, we used Higgins’ I² statistic, with an I² value greater than 50% indicating substantial heterogeneity. The I² value quantifies the proportion of variance (ranging from 0% to 100%) attributable to true effect sizes rather than sampling error. In cases of high heterogeneity, the random effects model was applied to estimate pooled ORs and MDs, while the fixed-effects model was used when heterogeneity was low.

## Results

### Maternal Outcomes

#### Incidence of Hypotension

Data on hypotension were pooled from nine studies (Study# 2, 3, 5, 6, 9-11, 13, 18) in Table 1 involving 640 parturients in the NE group and 643 in the PE group. No heterogeneity was observed (I² = 0%). The meta-analysis revealed no statistically significant difference in preventing or managing post-spinal hypotension between the two groups (OR = 1.17, 95% CI: 0.92 to 1.47), as presented in Figure 3.

**Fig. 3.**
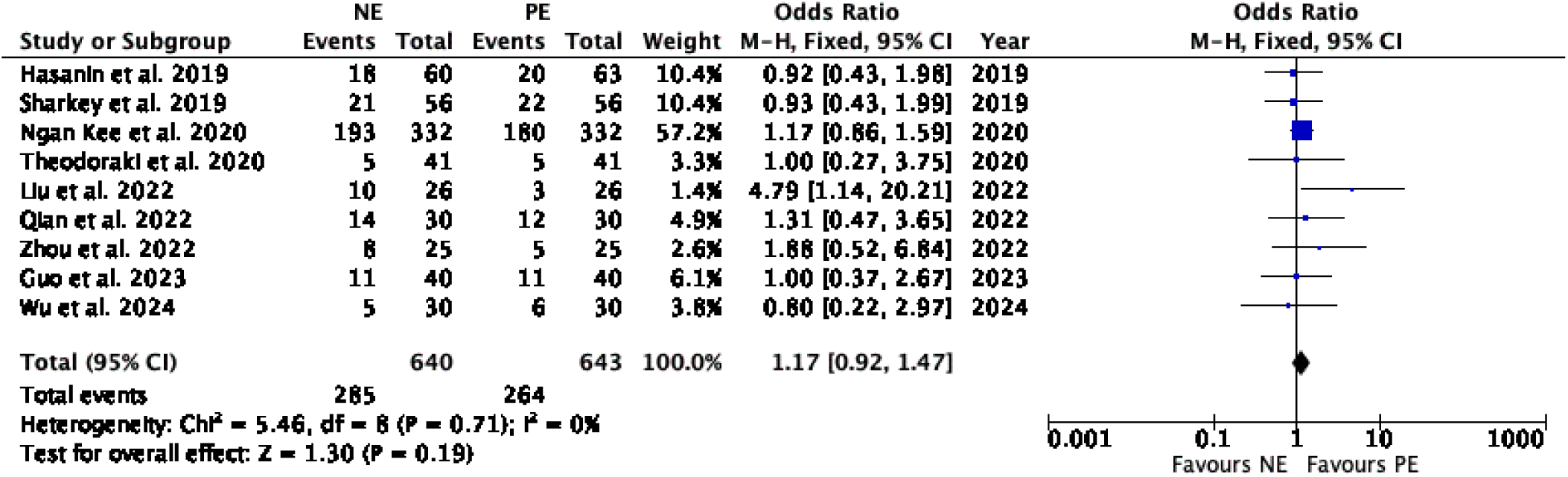
Incidence of hypotension between NE and PE (OR = 1.17 [CI: 0.92 to 1.47], I² = 0%, 9 trials, NE: N = 640, PE: N = 643). No heterogeneity was observed. The fixed-effects model revealed no statistically significant difference in hypotension incidence between the two groups.

#### Incidence of Bradycardia

Thirteen studies (Study# 1-7, 9, 11-13, 15, 18) in Table 1 involving 834 and 835 parturients in the norepinephrine (NE) and phenylephrine (PE) groups, respectively, reported on bradycardia. The pooled analysis showed moderate heterogeneity (I² = 44%). A fixed-effects model revealed that NE significantly reduced the incidence of bradycardia compared to PE (OR = 0.49, 95% CI: 0.38–0.62), as depicted in Figure 4.

**Fig. 4.**
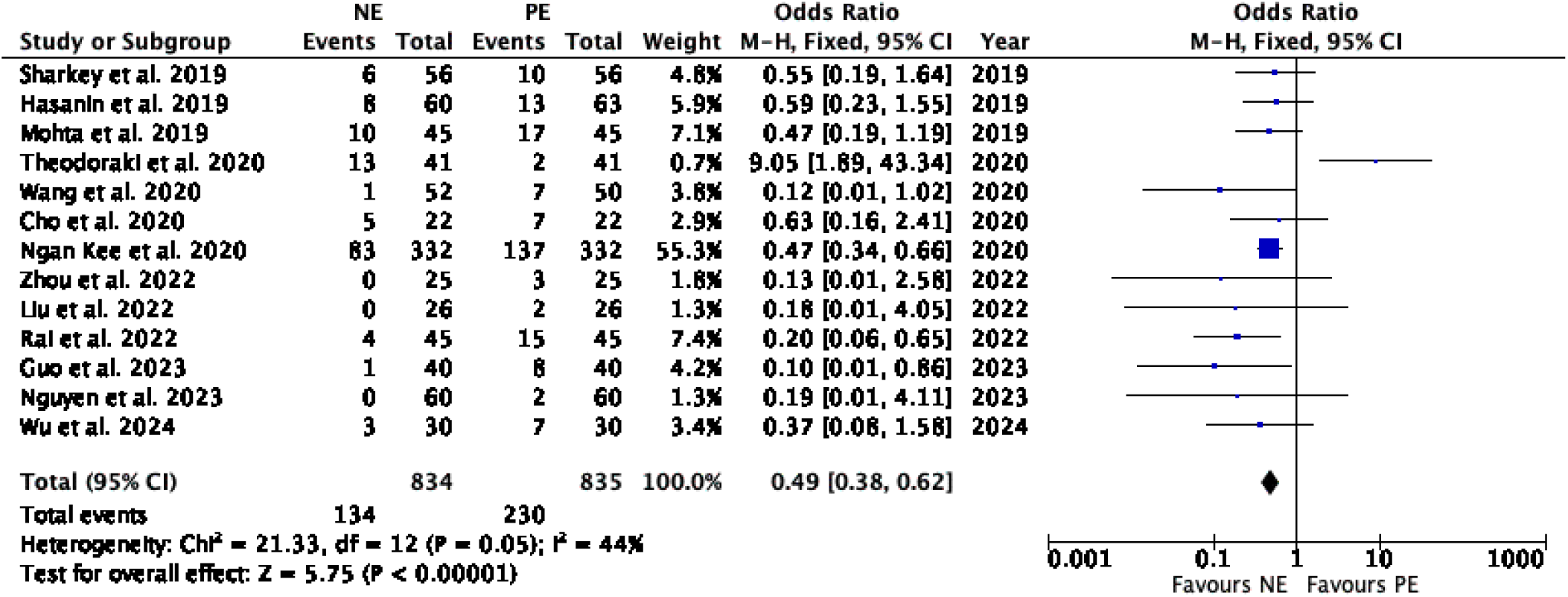
Incidence of bradycardia between NE and PE (OR = 0.49 [CI: 0.38 to 0.62], I² = 44%, 13 trials, NE: N = 834, PE: N = 835). The pooled analysis showed moderate heterogeneity. A fixed-effects model revealed that NE significantly reduced the incidence of bradycardia compared to PE.

#### Incidence of Dizziness

The incidence of dizziness was assessed in six studies (Study# 1, 5, 11, 12, 16, 18) in Table 1. involving 248 parturients in both NE and PE groups. There was no significant heterogeneity across the studies (I² = 28%). Pooled analysis using a fixed-effects model demonstrated no statistically significant difference in the incidence of dizziness between the NE and PE groups (OR = 0.62, 95% CI: 0.28 to 1.37), as shown in Figure 5.

**Fig. 5.**
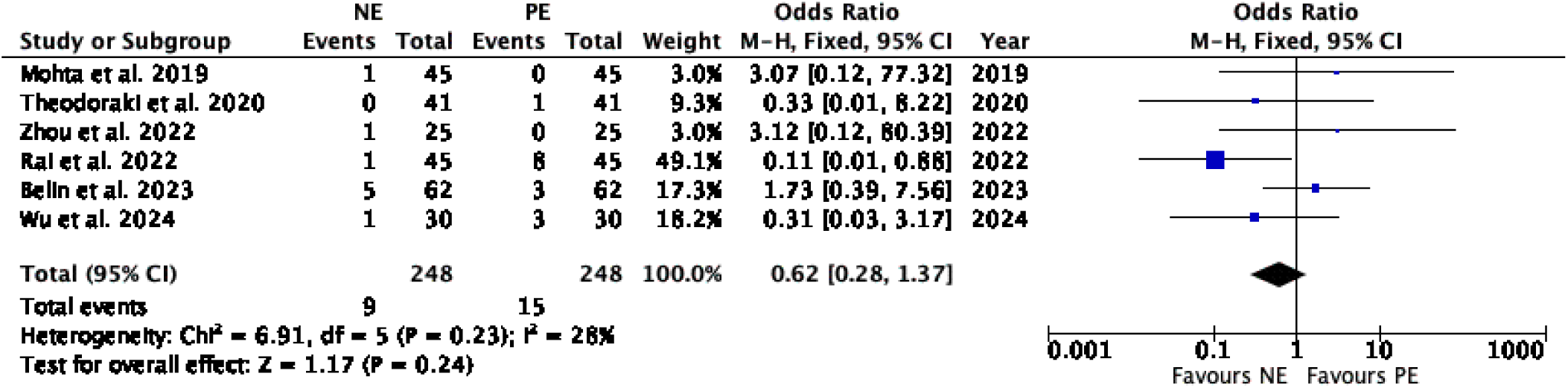
Incidence of dizziness between NE and PE (OR = 0.62 [CI: 0.28 to 1.37], I² = 28%, 6 trials, N = 248). There was no significant heterogeneity across the studies. Pooled analysis using a fixed-effects model demonstrated no statistically significant difference in the incidence of dizziness between the NE and PE groups.

#### Incidence of Reactive Hypertension

Reactive hypertension incidence was analyzed across six studies (Study# 3, 9-11, 13, 18) in Table 1 with 207 parturients in both NE and PE groups. There was no significant heterogeneity detected (I² = 6%). The pooled data showed a trend toward decreased incidence in the NE group, with an odds ratio of 0.58 (95% CI: 0.29 to 1.16), as presented in Figure 6.

**Fig. 6.**
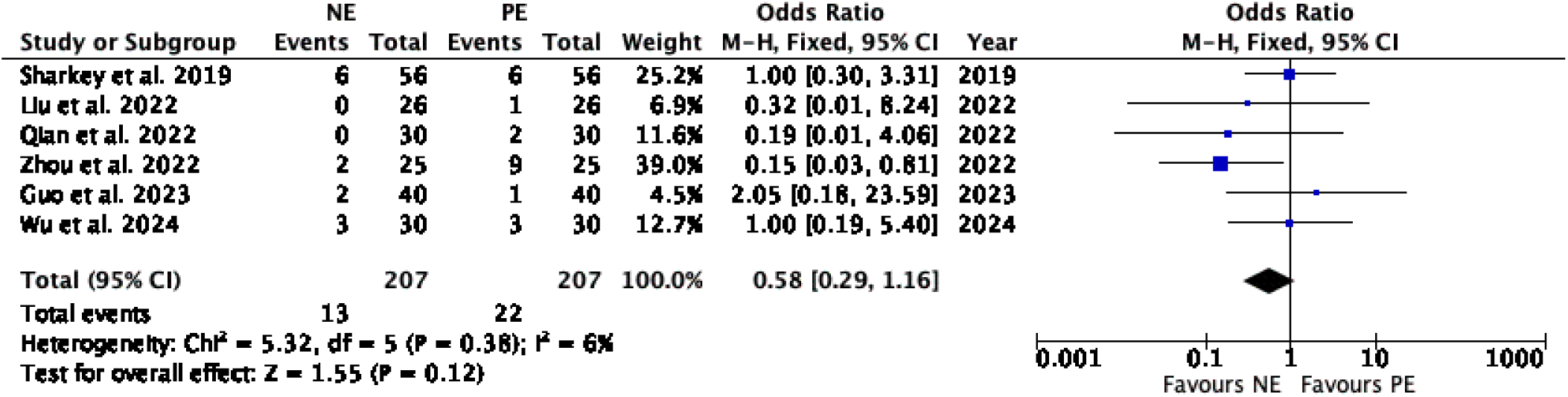
Incidence of reactive hypertension between NE and PE (OR = 0.58 [CI: 0.29 to 1.16], I² = 6%, 6 trials, N = 207). There was no significant heterogeneity detected (I² = 6%). The pooled data showed a trend toward decreased incidence in the NE group.

#### Incidence of Vomiting

Fourteen studies (Study# 1-4, 6-10, 12, 13, 15, 16, 18 in Table 1) with a total of 885 parturients in the NE group and 886 in the PE group were included to evaluate the incidence of vomiting. The analysis revealed no significant heterogeneity among the studies (I² = 0%), indicating consistent results across the data. Using a fixed-effects model, the findings showed no notable difference in vomiting incidence between the two groups (OR = 1.20, 95% CI: 0.91 to 1.57), as depicted in Figure S1.

### Neonatal Outcomes

#### Neonatal Birth Weight

In Figure 7, the eight trials (Study# 4-7, 10, 12, 15, 17 in Table 1) involving 629 parturients in the NE group and 628 in the PE group evaluated neonatal birth weight in kilograms. The analysis revealed moderate heterogeneity (I² = 41%). Using a fixed-effects model, the meta-analysis revealed a statistically significant difference in neonatal birth weight between the two groups. However, since both groups fall within the normal range of neonatal birth weight, this difference was considered clinically non-significant (MD = –0.07, 95%, CI: –0.09 to –0.05).

**Fig. 7.**
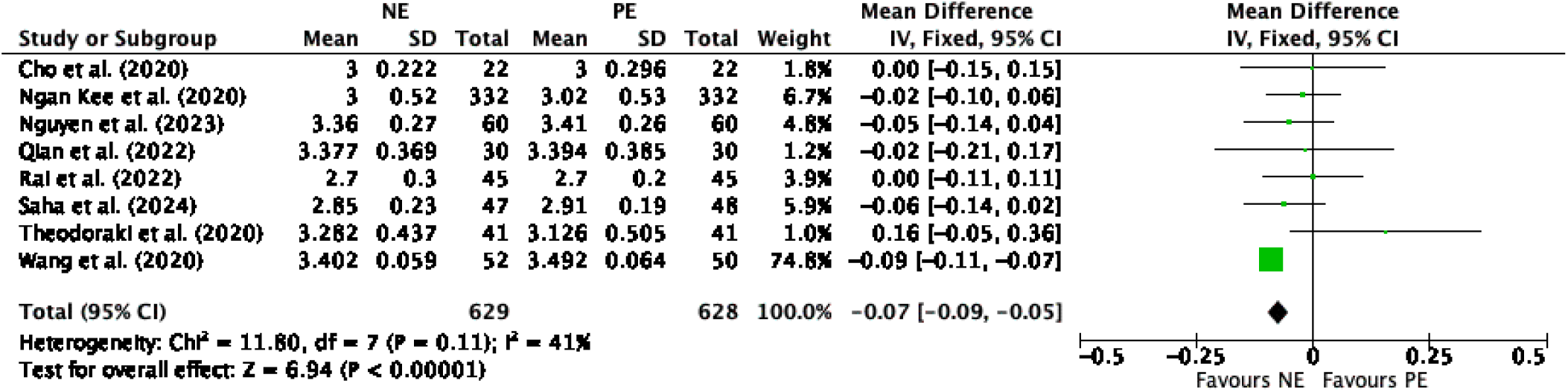
The mean difference in neonatal birth weight between NE and PE (OR = –0.07 [CI: –0.09 to –0.05], I² = 41%, 8 trials, NE: N = 629, PE: N = 628) indicated moderate heterogeneity. Using a fixed-effects model, the meta-analysis revealed a statistically significant difference in neonatal birth weight between the two groups. However, since both groups fall within the normal range of neonatal birth weight, this difference was considered clinically non-significant.

#### Umbilical Arterial Lactate Levels

Four studies (Study# 7, 9, 11, 15 in Table 1) involving 163 parturients in the NE group and 161 in the PE group evaluated umbilical arterial lactate levels. The analysis revealed significant heterogeneity (I² = 62%). Utilizing a random effects model, the results suggested a trend toward lower lactate levels in the NE group (MD = –0.11, 95% CI: –0.21 to 0.10), as seen in Figure 8.

**Fig. 8.**
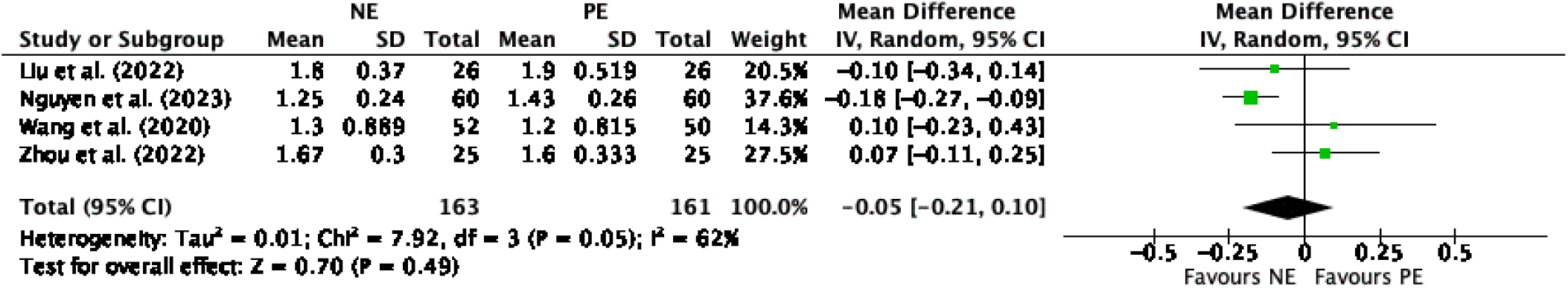
Mean difference in umbilical arterial lactate levels between NE and PE (OR = –0.05 [CI: –0.21 to 0.10], I² = 62%, 4 trials, NE: N = 163, PE: N = 161). The analysis revealed significant heterogeneity. Using a random effects model, the results indicated a trend toward lower lactate levels in the NE group.

#### Umbilical Vein Blood Gas pH

The pH values from blood gas analyses reported in the included studies were extracted, with data from nine studies (Study# 1, 3, 5-7, 9, 11, 12, 16 in Table 1) encompassing 684 parturients in the NE group and 682 in the PE group pooled to evaluate umbilical vein blood gas pH levels. Minimal heterogeneity was observed (I² = 12%). Analysis using a fixed-effects model demonstrated a trend toward a less acidic umbilical vein blood gas pH (MD = 0.00, 95% CI: 0.00 to 0.01), as illustrated in Figure 9.

**Fig. 9.**
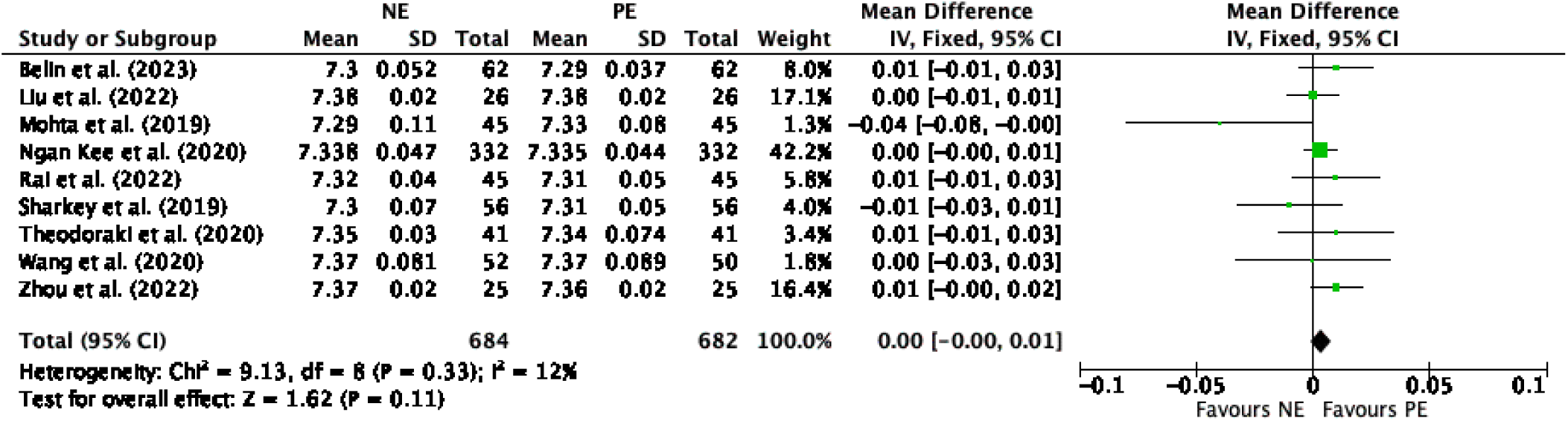
Mean difference in umbilical vein blood gas pH between NE and PE (OR = 0.00 [CI: 0.00 to 0.01], I² = 12%, 9 trials, NE: N = 684, PE: N = 682). Minimal heterogeneity was observed. Analysis using a fixed-effects model demonstrated a trend toward a less acidic umbilical vein blood gas pH.

#### Other Neonatal Outcomes

We analyzed additional outcome measures and found no significant differences in partial pressure of oxygen (PO₂) and carbon dioxide (PCO₂), bicarbonate (HCO₃⁻), and base excess in both the umbilical artery and vein, as illustrated in Figures S2–S7.

## Discussion

This study identified post-spinal hypotension as its primary outcome of interest. Hypotension is a common complication of spinal anesthesia during cesarean sections. This study found that the incidence of post-spinal hypotension in CS parturients is comparable between the two groups (Norepinephrine [NE] and Phenylephrine [PE]). Interestingly, NE significantly reduced the incidence of maternal bradycardia compared to PE. This concurs with the meta-analyses of Kumari et al. (2022) and Liu et al. (2022), which reported similar findings regarding the incidence of post-spinal hypotension, along with a reduced incidence of bradycardia with NE. We observed a tendency toward a reduction in other adverse maternal events, such as dizziness and hypotension, with NE compared to PE. Regarding neonatal outcomes, we found a statistically significant difference in neonatal birth weight between the two groups. However, since both groups fell within the normal range of neonatal birth weight, this difference was considered clinically non-significant. We also observed a trend toward lower umbilical arterial lactate levels with NE, as well as reduced fetal acidosis, as indicated by the umbilical vein blood gas pH analysis, compared to PE. No significant differences were observed in other neonatal outcomes. These findings update the previous meta-analyses by Kumari et al. (2022) and Liu et al. (2022), which compared NE and PE as vasopressors for preventing and managing post-spinal hypotension in cesarean section parturients.

Hypotension induced by spinal anesthesia occurs due to arterial and venous dilation caused by a sympathetic block, coupled with the paradoxical activation of cardioinhibitory receptors (Ferré et al., 2020). Šklebar et al. (2019) explain that the most notable pathophysiological mechanism by which spinal anesthesia causes hypotension is the rapid onset of sympatholysis. This occurs because parturients’ nerve fibers are more sensitive to local anesthetics. The increased incidence and severity of hypotension in pregnant women, compared to non-obstetric patients, are primarily attributed to their heightened sensitivity to local anesthetics and the aortocaval compression caused by the gravid uterus.

In a case report by Kalra and Hayaran (2011), a patient, after a subarachnoid block for lower uterine segment cesarean section (LSCS), showed bradycardia on ECG, which is a common finding following the administration of spinal anesthesia. Ferré et al. (2020) emphasize that bradycardia after spinal anesthesia should always be considered a warning sign of significant hemodynamic instability. This highlights the importance of addressing bradycardia with NE emerging as a promising candidate to alleviate this. They also noted that spinal anesthesia induces a sympathovagal imbalance in favor of parasympathetic tone, which leads to bradycardia and reduced blood pressure. In a review by Neal (2000), it was suggested that a reduced preload can trigger reflexes in response to changes in intracardiac volume or pacemaker receptors, contributing to bradycardia. A rapid decrease in left ventricular volume may cause severe bradycardia and asystole through paradoxical activation of the Bezold-Jarisch reflex, a rare but serious complication associated with spinal anesthesia. Kumari et al. (2022) and Liu et al. (2022) observed similar results in their studies, showing a significant difference between NE and PE. Kumari et al. (2022) suggested that NE’s effectiveness in reducing bradycardia could be attributed to its beta-adrenergic activation, which increases heart rate. In contrast, PE is an alpha-1 adrenergic receptor agonist, which acts as a vasoconstrictor and exerts an effect on total peripheral resistance (Richards et al. 2023). The pooled analysis of bradycardia from 13 trials showed a notable advantage for norepinephrine, indicating that bradycardia was less likely to occur with norepinephrine than with phenylephrine. This suggests that maternal hemodynamic instability associated with bradycardia can be more effectively prevented or treated with NE rather than PE.

On the other hand, while there was no significant difference between NE and PE in the maternal adverse event of dizziness, a trend suggests that norepinephrine may reduce its incidence. However, it is important to note that we cannot definitively conclude a clinically significant result for dizziness, as there is a lack of randomized controlled trials (RCTs) to support these findings. The pooled analysis found no significant difference between NE and PE in managing hypotension, which directly influences dizziness, nausea, and vomiting. This concurs with the meta-analysis by Liu et al. (2022). In addition to dizziness, reactive hypertension was found to have no significant difference between the two drugs, although NE may tend to reduce its incidence. A case-control study by Filho and Antunes (2020) found that hypertensive disorders are linked to higher rates of cesarean sections, as well as an increased likelihood of preterm birth, low birth weight infants, and fetal mortality. This suggests that parturients undergoing cesarean section are likely to experience hypertension. Our findings regarding NE’s impact on hypertension should also be considered in this context, suggesting that norepinephrine could potentially reduce the risk of hypertension. However, we must emphasize that we cannot establish a clinically significant outcome for hypertension due to the lack of supporting RCTs. While Liu et al. (2022) report a significant effect of NE in reducing the risk of hypertension, our study’s pooled analysis aligns with Kumari et al. (2022), where both NE and PE showed similar effects in parturients.

Regarding neonatal outcomes, our study found that neonatal birth weight tends to be lower with NE administration than with PE. However, despite this statistically significant difference, neonatal birth weights remained within the normal range. According to the World Health Organization (WHO), the average birth weight of a full-term male infant is approximately 3.3 kg (7 lbs 6 oz), while a full-term female infant weighs around 3.2 kg (7 lbs 2 oz) (Shehzad, 2011). It is important to note that for this outcome, the majority of the data analyzed in our study came from the randomized controlled trial conducted by Wang et al. (2020), which may have influenced the overall findings. This suggests that while NE may be associated with a lower neonatal birth weight, the difference is not necessarily clinically significant.

This study also found that NE may tend to reduce the risk of lactic acidosis compared to PE in neonatal outcomes. Neonatal lactate levels are important indicators of oxygen delivery and cellular function (Pirrone et al., 2012). Elevated lactate levels suggest a shift to anaerobic metabolism due to reduced oxygen supply, which leads to lactate accumulation, inadequate tissue perfusion, and potential hypoxia (Levy et al., 2000; Mikkelsen et al., 2009). If left unaddressed, this may result in lactic acidosis, increasing neonatal stress and the risk of adverse outcomes (Ganetzky & Cuddapah, 2017). Although Kumari et al. (2022) and Liu et al. (2022) did not report differences in lactate levels between NE and PE, our pooled analysis of umbilical arterial lactate levels from four trials revealed a trend toward a difference favoring NE, suggesting a possible improvement in neonatal oxygenation and perfusion.

Both NE and PE were comparable in maintaining a stable acid-base balance, with a trend toward NE resulting in less acidic umbilical vein blood gas levels, indicating that both drugs are similarly effective in supporting neonatal homeostasis. These results align with Kumari et al. (2022) and Liu et al. (2022), who also found no significant difference in pH between the two vasopressors. Other outcome measures, including partial pressure of oxygen (PO₂), carbon dioxide (PCO₂), bicarbonate (HCO₃⁻), and base excess in both the umbilical artery and vein, showed no significant differences between NE and PE.

## Conclusion

In this systematic review and meta-analysis of 18 randomized controlled trials comparing norepinephrine (NE) and phenylephrine (PE) for cesarean deliveries, NE demonstrated comparable efficacy to PE in preventing and managing post-spinal hypotension while significantly reducing the incidence of maternal bradycardia and showing a trend toward fewer adverse maternal events, such as dizziness and reactive hypertension. Neonatal outcomes revealed that, although neonates in the NE group had a statistically lower birth weight, this difference remained within the normal range, and there was a tendency toward lower umbilical arterial lactate levels and reduced acidosis as indicated by umbilical vein blood gas pH analysis. These findings, which update previous meta-analyses by Kumari et al. (2022) and Liu et al. (2022), support NE as a promising alternative to PE for mitigating both maternal and neonatal adverse events in the context of cesarean deliveries. To solidify the evidence supporting NE’s efficacy and safety as a first-line vasopressor during cesarean deliveries, we recommend further large-scale, multi-center randomized controlled trials. These trials should rigorously compare optimal dosing strategies (bolus vs. continuous infusion) and timing of administration (prophylactic vs. therapeutic), employing standardized protocols to minimize heterogeneity. Recognizing that inconsistent definitions and measurement techniques for hypotension contribute to heterogeneity across studies, future trials should adhere to universally accepted criteria (e.g., a decrease of ≥20% from baseline or an absolute value <90 mmHg for systolic blood pressure) and utilize validated measurement methods. These comprehensive investigations will be crucial for developing evidence-based clinical practice guidelines that optimize maternal and neonatal well-being during cesarean deliveries.

## Data Availability

All data from selected randomized controlled trials are available online at PubMed and Scopus

## Acknowledgments

R.D.G and P.K.Y would like to express their sincere gratitude to the Department of Science and Technology – Science Education Institute (DOST-SEI) for their financial support. We also acknowledge De La Salle University for providing the facilities and resources necessary for this study.

## Author contributions

**Conceptualization**, A.C., R.D.G., A.F.S., P.K.Y., J.T.C.; **Methodology**, A.C, R.D.G; **Validation**, A.F.S., P.K.Y., J.T.C.; **Formal analysis**, A.C., R.D.G.; **Investigation**, A.C, R.D.G; **Writing** – Original Draft, A.C, R.D.G; **Writing** – Review & Editing, A.F.S, P.K.Y, J.T.C.; **Funding Acquisition**, J.T.C.; **Data curation**, A.C, R.D.G; **Supervision**, A.F.S, P.K.Y, J.T.C.

