## Supplementary material for "Fetomaternal outcomes among cesarean section parturients administered with norepinephrine vs. phenylephrine for post-spinal anesthesia hypotension: A systematic review and meta-analysis": CAMBAGANADIN_SupplementalFiles_Fetomaternalou.html

 

### Maternal Outcomes

#### Fig. S1. Incidence of vomiting between NE and PE (OR = 1.20 [CI: 0.91 to 1.57], I² = 0%, 14 trials, NE: N = 885, PE: N = 886). The analysis revealed no significant heterogeneity among the studies. Using a fixed-effects model, the findings showed no notable difference in vomiting incidence between the two groups. ---

### Neonatal Outcomes

#### Fig. S2. Mean difference in umbilical arterial partial pressure of oxygen (PO2) between NE and PE (OR = 0.00 [CI: –0.91 to 0.90], I² = 62%, 15 trials, NE: N = 891, PE: N = 893). Substantial heterogeneity was observed. In the random effects model, the meta-analysis revealed no statistically significant difference in the umbilical arterial partial pressure oxygen (PO2) between the two groups. ---

#### Fig. S3. Mean difference in umbilical arterial partial pressure of carbon dioxide (PCO2) between NE and PE (OR = 0.03 [CI: –0.64 to 0.70], I² = 18%, 14 trials, NE: N = 851, PE: N = 853). Minimal heterogeneity was noted. Using a fixed-effects model, the meta-analysis revealed no statistically significant difference in umbilical arterial partial pressure of carbon dioxide (PCO2) between the two groups. ---

#### Fig. S4. Mean difference in umbilical venous partial pressure of carbon dioxide (PCO2) between NE and PE (OR = –0.31 [CI: –1.23 to 0.61], I² = 0%, 8 trials, NE: N = 352, PE: N = 350). No heterogeneity was observed. The meta-analysis indicated no statistically significant difference in umbilical venous partial pressure of carbon dioxide (PCO2) between the two groups as shown in the fixed-effects model. ---

#### Fig. S5. Mean difference in umbilical arterial bicarbonate (HCO3–) between NE and PE (OR = –0.10 [CI: –0.68 to 0.48], I² = 70%, 10 trials, NE: N = 442, PE: N = 444). Substantial heterogeneity was observed. The random effects model showed that the meta-analysis revealed no statistically significant difference in umbilical arterial bicarbonate (HCO3–) between the two groups. ---

#### Fig. S6. Mean difference in umbilical arterial base excess between NE and PE (OR = 0.07 [CI: –0.15 to 0.30], I² = 13%, 12 trials, NE: N = 778, PE: N = 777). The fixed-effects model used in the meta-analysis revealed no statistically significant difference in umbilical arterial base excess between the two groups. ---

#### Fig. S7. Mean difference in umbilical venous base excess between NE and PE (OR = 0.18 [CI: –0.29 to 0.66], I² = 70%, 9 trials, NE: N = 684, PE: N = 684). Substantial heterogeneity was observed. The meta-analysis revealed no statistically significant difference in umbilical venous base excess between NE and PE between the two groups as depicted through the random effects model.
