## Supplementary figures and images for "Fetomaternal outcomes among cesarean section parturients administered with norepinephrine vs. phenylephrine for post-spinal anesthesia hypotension: A systematic review and meta-analysis"

### image1.png

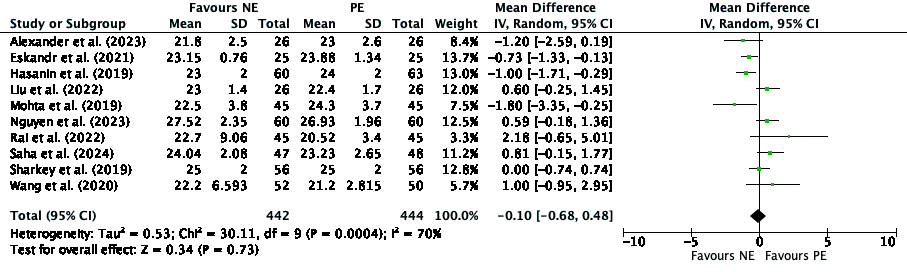

### image2.png

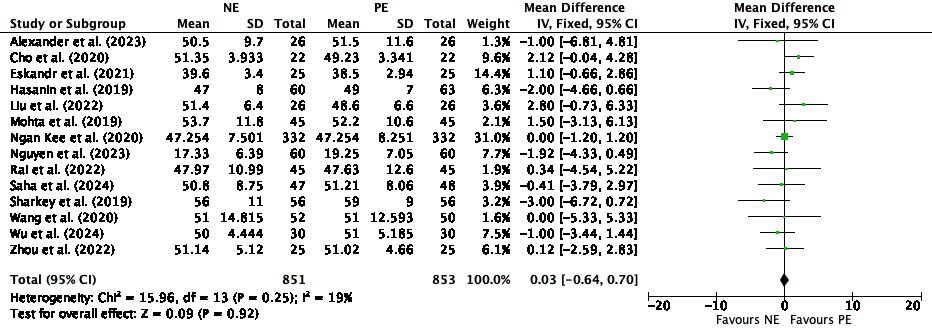

### image3.png

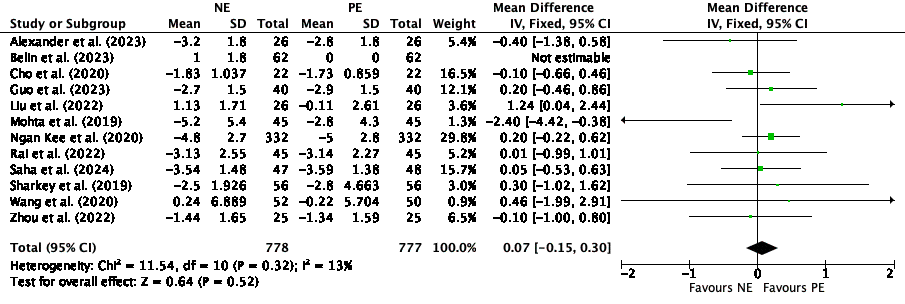

### image4.png

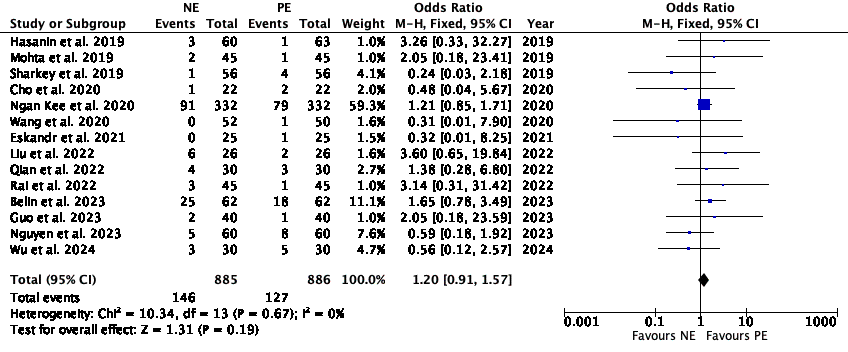

### image5.png

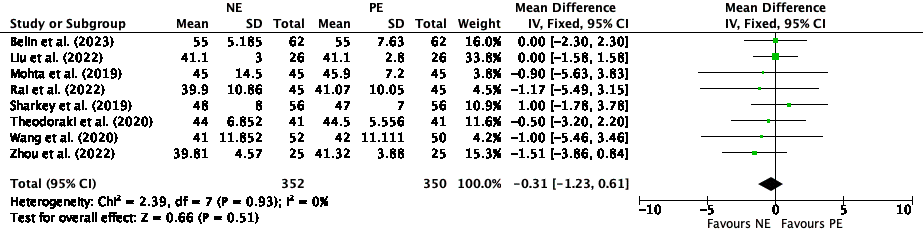

### image6.png

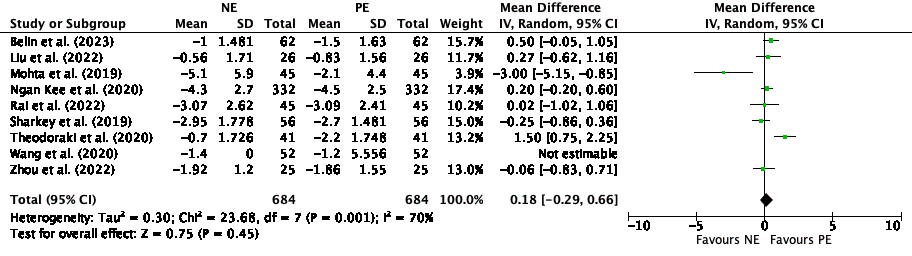

### image7.png

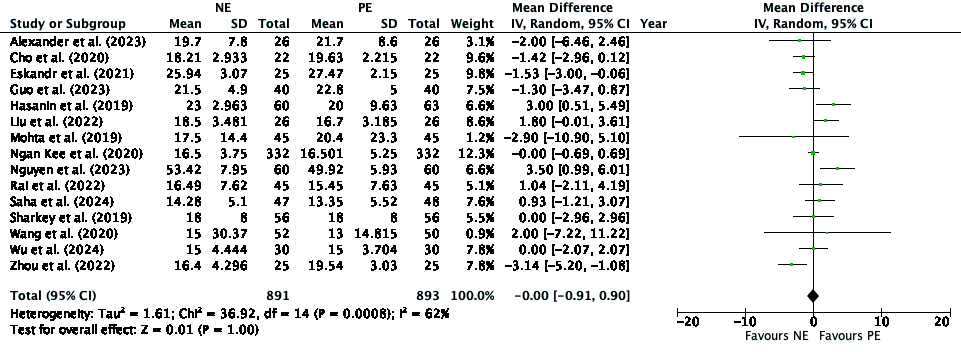
